# Mental Health, Substance Use, and Suicidal Ideation Among Unpaid Caregivers in the United States During the COVID-19 Pandemic: Relationships to Age, Race/Ethnicity, Employment, and Caregiver Intensity

**DOI:** 10.1101/2021.02.02.21251042

**Authors:** Mark É Czeisler, Alexandra Drane, Sarah S Winnay, Emily R Capodilupo, Charles A Czeisler, Shantha MW Rajaratnam, Mark E Howard

## Abstract

**Objectives:** To estimate the prevalence of unpaid caregiving during the coronavirus disease 2019 (COVID-19) pandemic, and to identify factors associated with adverse mental health symptoms, substance use, and suicidal ideation in this population, which provides critical support in health care systems by providing care to older adults and those with chronic conditions.

**Methods:** In June 2020, Internet-based surveys with questions about demographics, caregiving responsibilities, and mental health were administered to US adults aged ≥18 years. Demographic quota sampling and survey weighting to improve cross-sectional sample representativeness of age, gender, and race/ethnicity. Prevalence ratios for adverse mental health symptoms were estimated using multivariable Poisson regressions.

**Results:** Of 9,896 eligible invited adults, 5,412 (54.7%) completed surveys; 5,011 (92.6%) respondents met screening criteria and were analysed, including 1,362 (27.2%) caregivers. Caregivers had higher prevalences of adverse mental health symptoms than non-caregivers, including anxiety or depressive disorder symptoms (57.6% *vs* 21.5%, respectively, p<0.0001) having recently seriously considered suicide (33.4% *vs* 3.7%, p<0.0001). Symptoms were more common among caregivers who were young *vs* older adults (e.g., aged 18–24 *vs* ≥65 years, aPR 2.75, 95% CI 1.95–3.88, p<0.0001), Hispanic or Latino *vs* non-Hispanic White (1.14, 1.04–1.25, p=0.0044), living with *vs* without disabilities (1.18, 1.10–1.26, p<0.0001), and with moderate and high *vs* low Caregiver Intensity Index scores (2.31, 1.65–3.23; 2.81, 2.00–3.94; both p<0.0001). Suicidal ideation was more prevalent among non-Hispanic Black *vs* non-Hispanic White caregivers (1.48, 1.15–1.90, p=0.0022).

**Conclusions:** Caregivers, who accounted for one in four US adult respondents in this nationally representative sample, more commonly reported adverse mental health symptoms than non-caregivers. Increased visibility of and access to mental health care resources are urgently needed to address mental health challenges of caregiving.

## Introduction

The coronavirus disease 2019 (COVID-19) pandemic, caused by severe acute respiratory syndrome coronavirus 2 (SARS-CoV-2), has been associated with mental health challenges related to the morbidity and mortality caused by the disease and its mitigation. Early studies have documented elevated levels of adverse mental health symptoms in the United States^1,2^ and around the globe^3,4^ compared with previous years. Young adults and unpaid caregivers for adults (caregivers) are among highly affected populations.

A pre-pandemic meta-analysis found that caregivers, who perform activities such as assisting others with activities of daily living and medical tasks, experienced higher levels of depression and perceived stress and lower levels of general well-being than non-caregivers.^5^ Subsequent studies have characterized an association between subjective caregiver burden and depressive symptoms,^6^ which in some cases limited provision of care.^7^

During June 2020, caregivers reported significantly higher prevalence of adverse mental and behavioural health symptoms than non-caregivers, including symptoms of an anxiety disorder, depressive disorder, or COVID-19-related trauma- and stressor-related disorder (TSRD), having started or increased substance use to cope with the pandemic, and suicidal ideation.^1^ A study of 1,459 paediatric and adult brain tumour patients and 530 caregivers in 33 countries found that caregivers were significantly more anxious than patients, and 42.8% of caregivers felt that their caregiver burden has significantly increased during the pandemic.^8^

Caregivers represent a significant demographic in the United States. In 2020, the pre-pandemic estimated prevalence of caregivers was 19.2% of adults aged ≥18 years, or 47.9 million Americans.^9^ This estimate represents an increase in caregivers of more than eight million compared to 2015.^10^ Some people may have taken up unplanned caregiving roles during the pandemic due to mobility restrictions related to community mitigation activities designed to reduce potential exposure to SARS-CoV-2 for older adults. Moreover, others who were providing care before the pandemic may have faced barriers and disruptions to their routines and livelihood. Both scenarios require caregivers to make sacrifices to care for others during a time when their own lives may have been disrupted.

Addressing the needs of the disproportionately affected population of caregivers is critically important for the health and well-being of caregivers and the persons for whom they provide care. To effectively address these needs during the COVID-19 pandemic and afterwards, studies are needed to determine the prevalence and characteristics of caregivers, and to identify stressors that may be targets for support systems and prevention and intervention efforts. This study aims to estimate the prevalence and characteristics of US caregivers during the COVID-19 pandemic, and to evaluate factors associated with differences in mental and behavioural health symptoms.

## Methods

### Study design and participants

To assess mental and behavioural health among adults aged ≥18 years with residence in the United States who had provided unpaid care for adults during the COVID-19 pandemic, we conducted a cross-sectional analysis of an Internet-based survey study conducted during June 24–30, 2020 for The COVID-19 Outbreak Public Evaluation (COPE) Initiative. Surveys were administered by Qualtrics, LLC (Provo, Utah, and Seattle, Washington, US), a commercial survey company with a network of participant pools consisting of hundreds of suppliers. Further details on Qualtrics recruitment and methodology are provided in the appendix (p 1).

Participants included both first-time respondents and respondents who had completed a related survey during April 2–8, May 5–12, 2020, or both intervals. Demographic quota sampling was used to recruit respondents based on population estimates for age, gender, race, and ethnicity based on the 2010 US Census. Potential respondents likely to qualify based on demographic characteristics listed in their Qualtrics panellist profile were targeted during recruitment; demographic questions were then included in the survey to determine their eligibility. Potential respondents received invitations and could opt to participate by activating a survey link directing them to the participant information and consent page preceding the survey. Ineligible respondents who did not meet inclusion criteria (e.g., age <18 years, not a US resident) or exceeded set quotas (i.e., maximum demographic characteristic quota already met) were not empanelled in the survey.

### Survey instrument

The survey instruments included individual questions, validated questionnaires, and COVID-19-specific questionnaires used to assess respondent attitudes, behaviours, and beliefs related to COVID-19 and its mitigation, along with mental and behavioural health consequences of the COVID-19 pandemic.

Demographic variables included gender, categorized age, combined race/ethnicity, disability status, marital status, household occupancy, 2019 household income, US Census region, urban/rural classification using self-reported ZIP codes, employment status, and, among employed respondents, self-identified essential worker status and weekly paid work hours. Caregiving variables included the method by which caregivers provided care (in-person in-home only; in-person out-of-home only; virtually only; and both in-person and virtually), the person for whom they were providing care, weekly unpaid caregiving hours, caregiver experience in months, and caregiving intensity assessed using the 12- or 14-item ARCHANGELS Caregiver Intensity Index (CII; see appendix (p 1) for additional details), which is composed of three subscales: Caregiver Load based on four items (situation stability, impact on expenses, family strife, and preparedness), Caregiver Impacts based on four items (emotional state, work, personal time, and stress), and Caregiver Buffers based on six items (support, insurance knowledge, self-efficacy, financial knowledge, sense of purpose, and employer support). Caregivers who were also employed completed all 14 items, while those who were not employed completed all items except for the work and employer support items. The sum of items in each subscale is normalized from 0–100, and the normalized sum of the three subscales is used to categorize total CII scores as Low (0–25), Moderate (26–55), or High (≥56).

Symptoms of anxiety or depression were assessed via the four-item Patient Health Questionnaire (PHQ-4), a clinically validated screening instrument.^11^ Symptoms of COVID-19-related trauma- and stressor-related disorder (COVID-19 TSRD) were assessed via the six-item Impact of Event Scale (IES-6) to screen for overlapping symptoms of posttraumatic stress disorder (PTSD), acute stress disorder (ASD), and adjustment disorders (ADs).^12^ Respondents also reported whether they had started or increased substance use, (e.g., alcohol, drugs) to cope with stress or emotions related to COVID-19, or if they had seriously considered trying to kill themselves in the prior 30 days. See appendix (pp 1–2) for additional details.

### Quality screening

All surveys underwent Qualtrics, LLC standard data quality screening procedures, and a secondary cleaning conducted by the investigators; see appendix (p 2). Respondents who failed an attention or speed check, along with any responses that failed data quality screening procedures, were excluded from the analysis.

### Statistical analysis

Iterative proportional fitting and weight trimming (0.3≤weight≤3.0) were employed to improve the cross-sectional sample representativeness of the 2010 US population by age, gender, and combined race/ethnicity (appendix p 2). The statistical analyses were completed in four phases. The first phase included bivariate analyses and described demographic characteristics of caregivers and non-caregivers, as well as mental and behavioural health, overall and by demographic variables, among caregivers and non-caregivers. The second phase described mental and behavioural health symptoms among caregivers, overall and by both demographic variables and caregiving characteristics. For comparisons between (demographics, group prevalences of adverse mental and behavioural health symptoms) and among (prevalences of adverse mental or behavioural health symptoms by demographics) caregivers and non-caregivers, Rao-Scott adjusted Pearson chi-squared tests were used to test for differences in observed and expected frequencies among groups by characteristic with a Bonferroni adjustment and evaluated at a significance level of α = 0.05. The third phase included adjusted prevalence ratios (aPRs) and 95% confidence intervals (95% CIs) for adverse mental and behavioural health symptoms among caregivers estimated using Poisson regressions with robust standard errors evaluated at a significance level of α = 0.05. Finally, in the fourth phase, non-parametric Spearman correlations were calculated between individual CII items and mental and behavioural health measures to assess the relative association of each item with adverse mental and behavioural health. All statistical analyses were conducted using Python (version 3.7.8; Python Software Foundation) and using R software (version 4.0.2; The R Foundation) with the R survey package (version 3.29).

### Study approval and informed consent

The Monash University Human Research Ethics Committee reviewed and approved the study protocol (ID #24036). This activity was also reviewed by CDC and was conducted consistent with applicable federal law and CDC policy (45 CFR part 46, 21 CFR part 56; 42 USC Sect 241(d); 5 USC Sect 552a; 44 USC Sect 3501 et seq). All participants provided informed electronic consent prior to study commencement. Investigators received anonymised responses.

## Results

Of 9,896 eligible invited adults, 5,412 (54.7%) completed Internet-based surveys during June 24–30, 2020, including 3,638 (68.1%) first-time respondents and 1,729 (31.9%) respondents who first completed a survey for The COPE Initiative during April 2–8, 2020. Among the 5,412 respondents, 5,011 (92.6%) met secondary screening criteria and were included in this analysis (Figure 1). These 5,011 respondents included 1,362 (27.2%) caregivers and 3,649 (72.8%) non-caregivers (Table 1). There was not a significant difference in caregiver status by gender or 2019 household income, though compared to non-caregivers, caregivers were significantly more commonly of young age (e.g., 18–24 years=26.6% *vs* 8.0%, respectively, group p<0.0001) and either Black or Hispanic race/ethnicity (Black=18.8% *vs* 9.7%; Hispanic=29.0% *vs* 11.6%, group p<0.0001). White respondents accounted for 44.5% of caregivers and 70.8% of non-caregivers. Caregivers also more commonly reported living with a disability than not (37.9% *vs* 17.0%, p<0.0001), and, among employed caregivers, essential than nonessential worker status (73.7% *vs* 47.8%, p<0.0001) (Table 1).

**Table 1.**
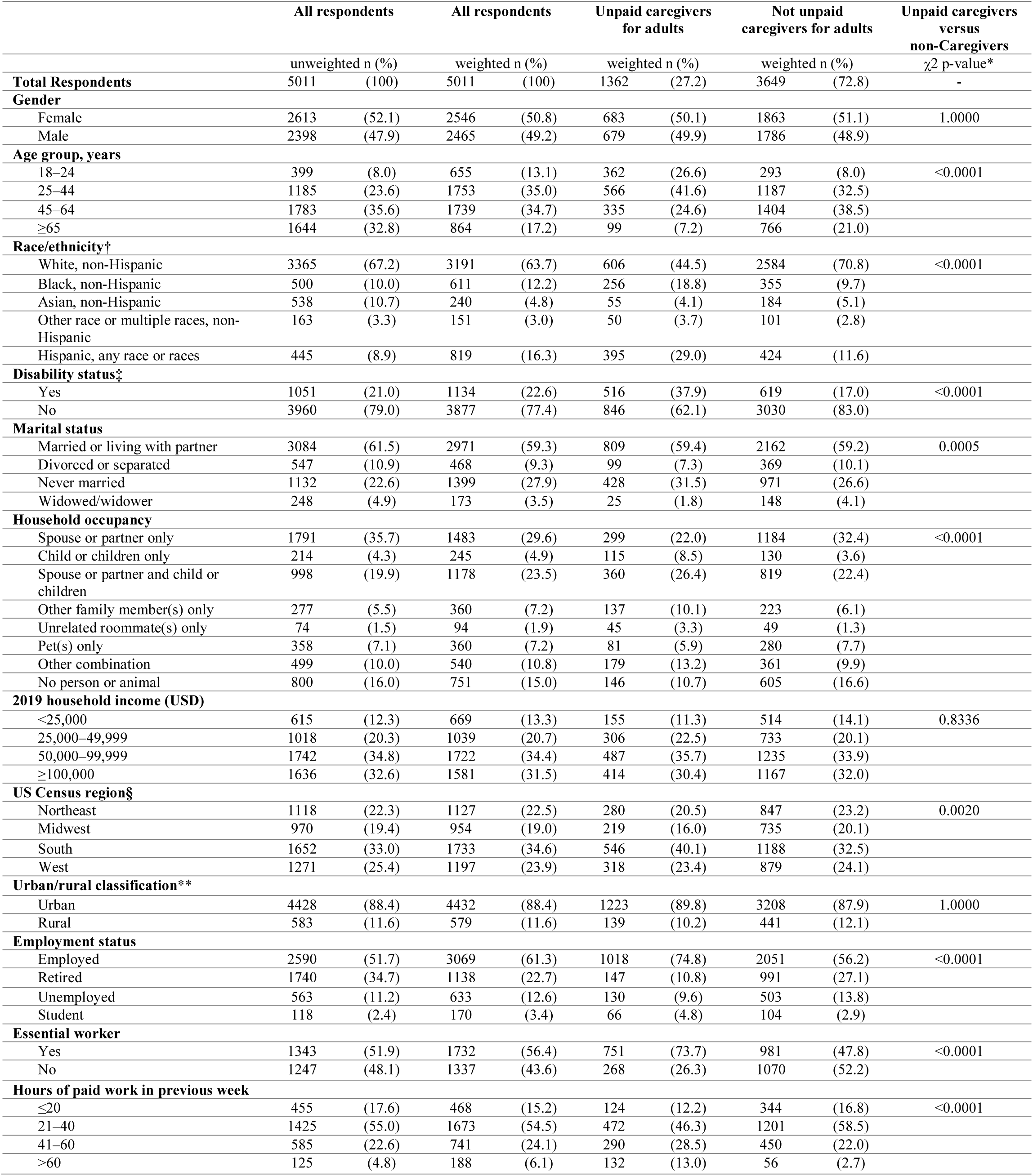
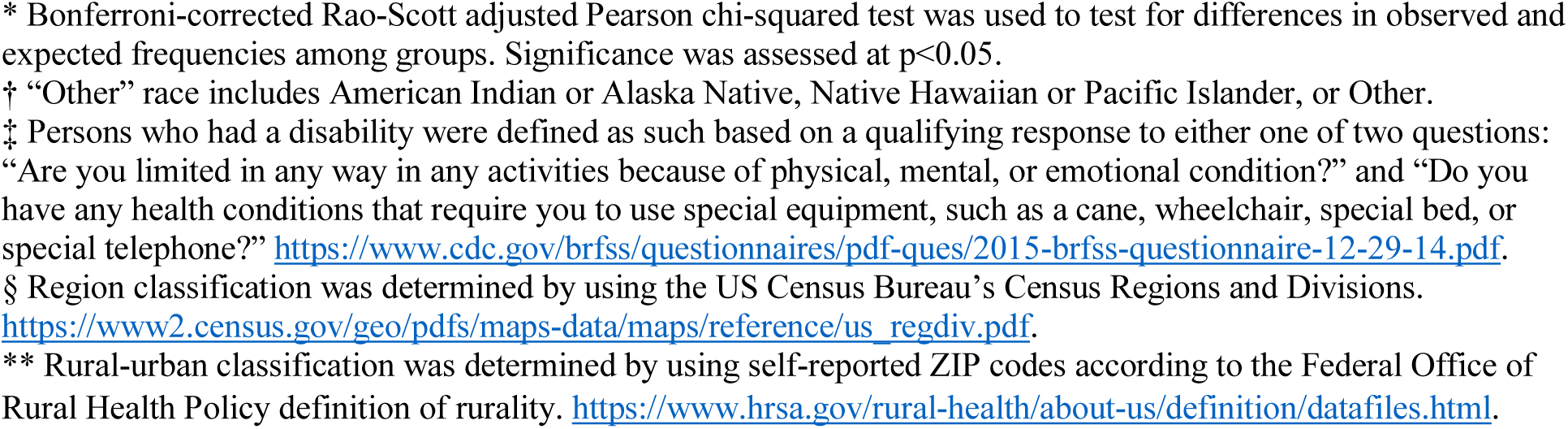
Respondent Characteristics by Caregiver Status.

|  | All respondents |  | All respondents |  | Unpaid caregivers for adults |  | Not unpaid caregivers for adults |  | Unpaid caregivers versus non-Caregivers |
| --- | --- | --- | --- | --- | --- | --- | --- | --- | --- |
| | unweighted n (%) | | weighted n (%) | | weighted n (%) | | weighted n (%) | | $\chi^2$ p-value* |
| **Total Respondents** | 5011 | (100) | 5011 | (100) | 1362 | (27.2) | 3649 | (72.8) | - |
| **Gender** |  |  |  |  |  |  |  |  |  |
| Female | 2613 | (52.1) | 2546 | (50.8) | 683 | (50.1) | 1863 | (51.1) | 1.0000 |
| Male | 2398 | (47.9) | 2465 | (49.2) | 679 | (49.9) | 1786 | (48.9) |  |
| **Age group, years** |  |  |  |  |  |  |  |  |  |
| 18–24 | 399 | (8.0) | 655 | (13.1) | 362 | (26.6) | 293 | (8.0) | <0.0001 |
| 25–44 | 1185 | (23.6) | 1753 | (35.0) | 566 | (41.6) | 1187 | (32.5) |  |
| 45–64 | 1783 | (35.6) | 1739 | (34.7) | 335 | (24.6) | 1404 | (38.5) |  |
| ≥65 | 1644 | (32.8) | 864 | (17.2) | 99 | (7.2) | 766 | (21.0) |  |
| **Race/ethnicity†** |  |  |  |  |  |  |  |  |  |
| White, non-Hispanic | 3365 | (67.2) | 3191 | (63.7) | 606 | (44.5) | 2584 | (70.8) | <0.0001 |
| Black, non-Hispanic | 500 | (10.0) | 611 | (12.2) | 256 | (18.8) | 355 | (9.7) |  |
| Asian, non-Hispanic | 538 | (10.7) | 240 | (4.8) | 55 | (4.1) | 184 | (5.1) |  |
| Other race or multiple races, non-Hispanic | 163 | (3.3) | 151 | (3.0) | 50 | (3.7) | 101 | (2.8) |  |
| Hispanic, any race or races | 445 | (8.9) | 819 | (16.3) | 395 | (29.0) | 424 | (11.6) |  |
| **Disability status‡** |  |  |  |  |  |  |  |  |  |
| Yes | 1051 | (21.0) | 1134 | (22.6) | 516 | (37.9) | 619 | (17.0) | <0.0001 |
| No | 3960 | (79.0) | 3877 | (77.4) | 846 | (62.1) | 3030 | (83.0) |  |
| **Marital status** |  |  |  |  |  |  |  |  |  |
| Married or living with partner | 3084 | (61.5) | 2971 | (59.3) | 809 | (59.4) | 2162 | (59.2) | 0.0005 |
| Divorced or separated | 547 | (10.9) | 468 | (9.3) | 99 | (7.3) | 369 | (10.1) |  |
| Never married | 1132 | (22.6) | 1399 | (27.9) | 428 | (31.5) | 971 | (26.6) |  |
| Widowed/widower | 248 | (4.9) | 173 | (3.5) | 25 | (1.8) | 148 | (4.1) |  |
| **Household occupancy** |  |  |  |  |  |  |  |  |  |
| Spouse or partner only | 1791 | (35.7) | 1483 | (29.6) | 299 | (22.0) | 1184 | (32.4) | <0.0001 |
| Child or children only | 214 | (4.3) | 245 | (4.9) | 115 | (8.5) | 130 | (3.6) |  |
| Spouse or partner and child or children | 998 | (19.9) | 1178 | (23.5) | 360 | (26.4) | 819 | (22.4) |  |
| Other family member(s) only | 277 | (5.5) | 360 | (7.2) | 137 | (10.1) | 223 | (6.1) |  |
| Unrelated roommate(s) only | 74 | (1.5) | 94 | (1.9) | 45 | (3.3) | 49 | (1.3) |  |
| Pet(s) only | 358 | (7.1) | 360 | (7.2) | 81 | (5.9) | 280 | (7.7) |  |
| Other combination | 499 | (10.0) | 540 | (10.8) | 179 | (13.2) | 361 | (9.9) |  |
| No person or animal | 800 | (16.0) | 751 | (15.0) | 146 | (10.7) | 605 | (16.6) |  |
| **2019 household income (USD)** |  |  |  |  |  |  |  |  |  |
| <25,000 | 615 | (12.3) | 669 | (13.3) | 155 | (11.3) | 514 | (14.1) | 0.8336 |
| 25,000–49,999 | 1018 | (20.3) | 1039 | (20.7) | 306 | (22.5) | 733 | (20.1) |  |
| 50,000–99,999 | 1742 | (34.8) | 1722 | (34.4) | 487 | (35.7) | 1235 | (33.9) |  |
| ≥100,000 | 1636 | (32.6) | 1581 | (31.5) | 414 | (30.4) | 1167 | (32.0) |  |
| **US Census region§** |  |  |  |  |  |  |  |  |  |
| Northeast | 1118 | (22.3) | 1127 | (22.5) | 280 | (20.5) | 847 | (23.2) | 0.0020 |
| Midwest | 970 | (19.4) | 954 | (19.0) | 219 | (16.0) | 735 | (20.1) |  |
| South | 1652 | (33.0) | 1733 | (34.6) | 546 | (40.1) | 1188 | (32.5) |  |
| West | 1271 | (25.4) | 1197 | (23.9) | 318 | (23.4) | 879 | (24.1) |  |
| **Urban/rural classification\*\*** |  |  |  |  |  |  |  |  |  |
| Urban | 4428 | (88.4) | 4432 | (88.4) | 1223 | (89.8) | 3208 | (87.9) | 1.0000 |
| Rural | 583 | (11.6) | 579 | (11.6) | 139 | (10.2) | 441 | (12.1) |  |
| **Employment status** |  |  |  |  |  |  |  |  |  |
| Employed | 2590 | (51.7) | 3069 | (61.3) | 1018 | (74.8) | 2051 | (56.2) | <0.0001 |
| Retired | 1740 | (34.7) | 1138 | (22.7) | 147 | (10.8) | 991 | (27.1) |  |
| Unemployed | 563 | (11.2) | 633 | (12.6) | 130 | (9.6) | 503 | (13.8) |  |
| Student | 118 | (2.4) | 170 | (3.4) | 66 | (4.8) | 104 | (2.9) |  |
| **Essential worker** |  |  |  |  |  |  |  |  |  |
| Yes | 1343 | (51.9) | 1732 | (56.4) | 751 | (73.7) | 981 | (47.8) | <0.0001 |
| No | 1247 | (48.1) | 1337 | (43.6) | 268 | (26.3) | 1070 | (52.2) |  |
| **Hours of paid work in previous week** |  |  |  |  |  |  |  |  |  |
| ≤20 | 455 | (17.6) | 468 | (15.2) | 124 | (12.2) | 344 | (16.8) | <0.0001 |
| 21–40 | 1425 | (55.0) | 1673 | (54.5) | 472 | (46.3) | 1201 | (58.5) |  |
| 41–60 | 585 | (22.6) | 741 | (24.1) | 290 | (28.5) | 450 | (22.0) |  |
| >60 | 125 | (4.8) | 188 | (6.1) | 132 | (13.0) | 56 | (2.7) |  |

\* Bonferroni-corrected Rao-Scott adjusted Pearson chi-squared test was used to test for differences in observed and expected frequencies among groups. Significance was assessed at $p < 0.05$ .
† “Other” race includes American Indian or Alaska Native, Native Hawaiian or Pacific Islander, or Other.
‡ Persons who had a disability were defined as such based on a qualifying response to either one of two questions: “Are you limited in any way in any activities because of physical, mental, or emotional condition?” and “Do you have any health conditions that require you to use special equipment, such as a cane, wheelchair, special bed, or special telephone?” https://www.cdc.gov/brfss/questionnaires/pdf-ques/2015-brfss-questionnaire-12-29-14.pdf.
§ Region classification was determined by using the US Census Bureau’s Census Regions and Divisions. https://www2.census.gov/geo/pdfs/maps-data/maps/reference/us_regdiv.pdf.
\*\* Rural-urban classification was determined by using self-reported ZIP codes according to the Federal Office of Rural Health Policy definition of rurality. https://www.hrsa.gov/rural-health/about-us/definition/datafiles.html.

**Figure 1.**
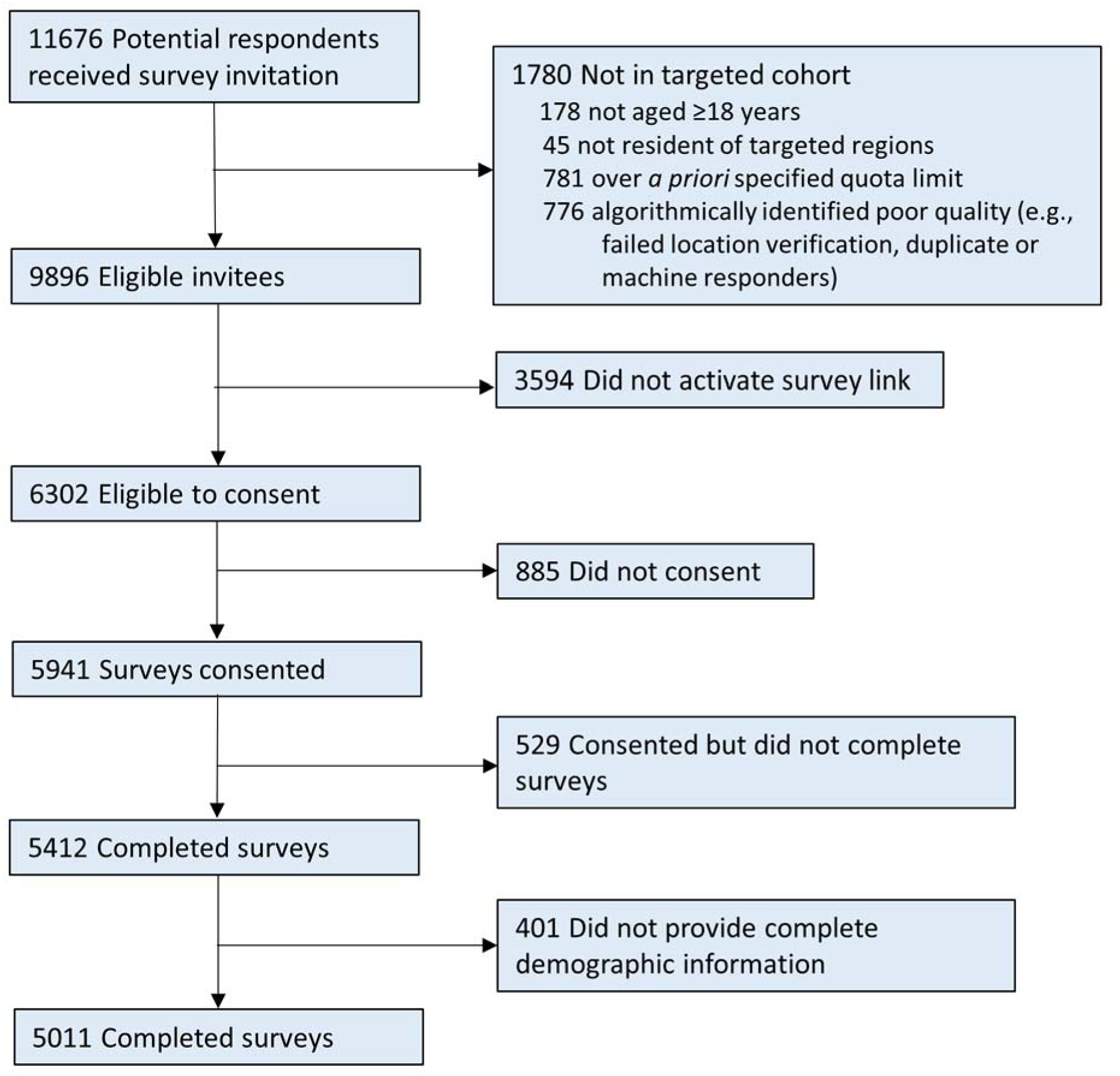
Flow of Survey Respondents.

Adverse mental and behavioural health symptoms were more prevalent among caregivers than non-caregivers (symptoms of anxiety or depressive disorder=57.6% *vs* 21.5%, respectively; symptoms of a COVID-19-related TSRD=49.0% *vs* 17.9%; having started or increased substance use to cope with the pandemic=35.0% *vs* 6.3%; suicidal ideation=33.4% *vs* 3.7%; at least one of these symptoms=69.6% *vs* 31.0%; all p<0.0001) (Tables 2,3).

**Table 2.**
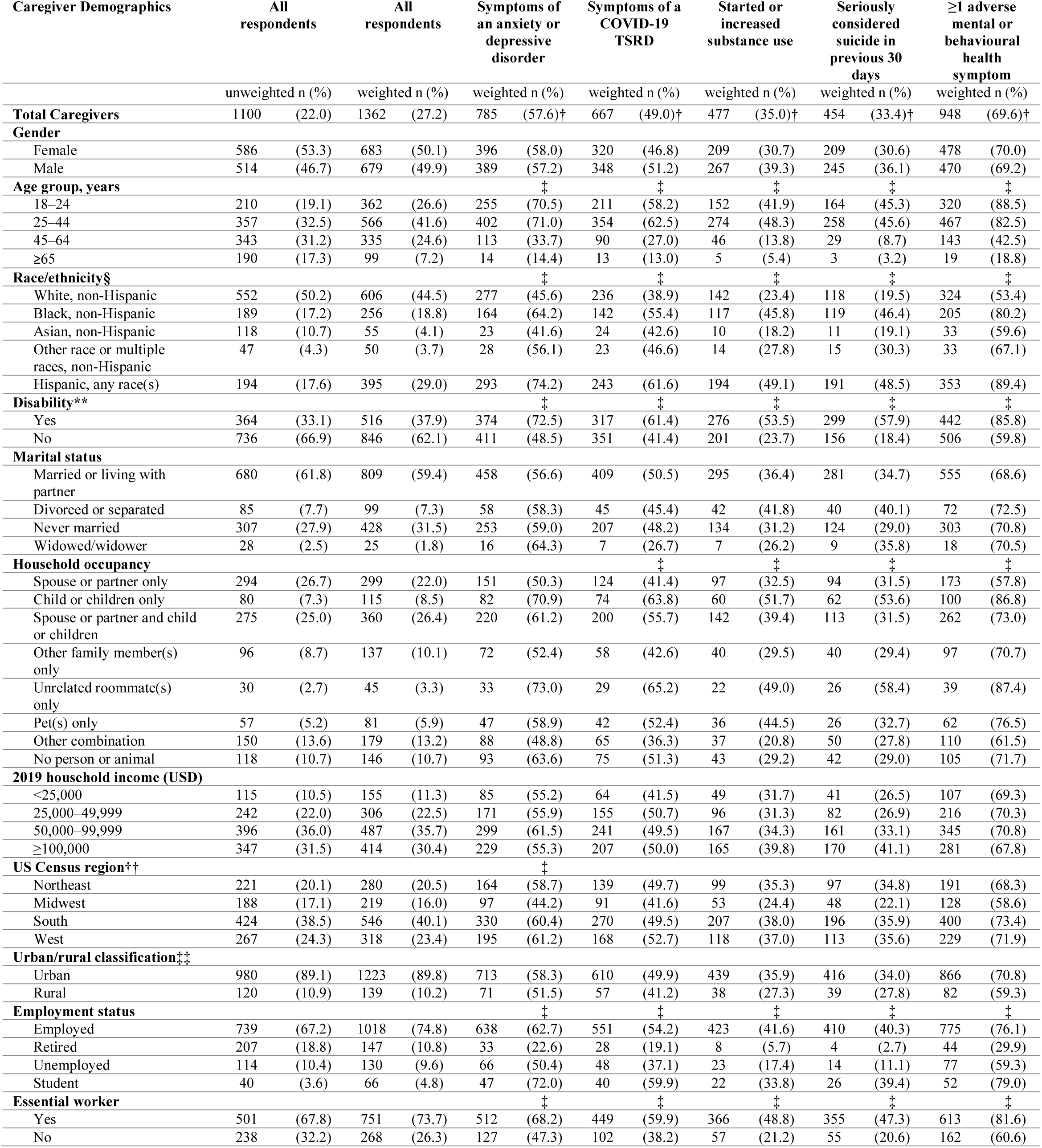
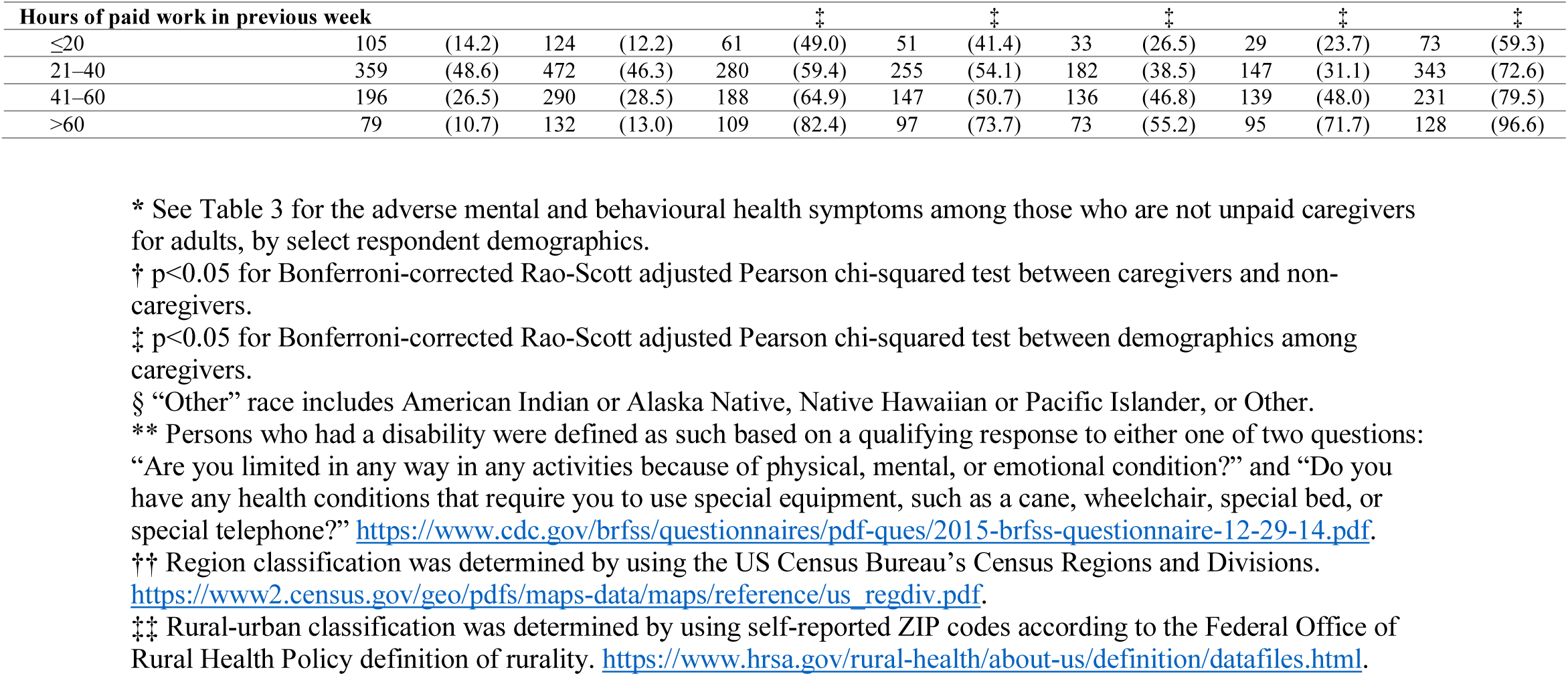
Adverse Mental and Behavioural Health Symptoms Among Unpaid Caregivers for Adults During June 24–30, 2020, by Select Respondent Demographics*.

**Table 3.** Adverse Mental and Behavioural Health Symptoms Among Those Who Are Not Unpaid Caregivers for Adults During June 24–30, 2020, by Select Respondent Demographics*.

| Non-Caregiver Demographics | All respondents |  | All respondents |  | Symptoms of an anxiety or depressive disorder |  | Symptoms of a COVID-19 TSRD |  | Started or increased substance use |  | Seriously considered suicide in previous 30 days |  | ≥1 adverse mental or behavioural health symptom |  |
| --- | --- | --- | --- | --- | --- | --- | --- | --- | --- | --- | --- | --- | --- | --- |
|  | unweighted n (%) |  | weighted n (%) |  | weighted n (%) |  | weighted n (%) |  | weighted n (%) |  | weighted n (%) |  | weighted n (%) |  |
| **Total Non-Caregivers** | 3911 | (78.0) | 3649 | (72.8) | 785 | (21.5)† | 653 | (17.9)† | 231 | (6.3)† | 135 | (3.7)† | 1130 | (31.0)† |
| **Gender** |  |  |  |  |  | ‡ |  |  |  |  |  |  |  | ‡ |
| Female | 2027 | (51.8) | 1863 | (51.1) | 475 | (25.5) | 355 | (19.1) | 134 | (7.2) | 76 | (4.1) | 655 | (35.2) |
| Male | 1884 | (48.2) | 1786 | (48.9) | 310 | (17.4) | 298 | (16.7) | 97 | (5.4) | 60 | (3.3) | 475 | (26.6) |
| **Age group, years** |  |  |  |  |  | ‡ |  |  |  |  |  |  |  | ‡ |
| 18–24 | 189 | (4.8) | 293 | (8.0) | 161 | (54.9) | 116 | (39.7) | 35 | (11.8) | 41 | (13.9) | 198 | (67.6) |
| 25–44 | 828 | (21.2) | 1187 | (32.5) | 334 | (28.1) | 303 | (25.5) | 105 | (8.9) | 56 | (4.7) | 474 | (39.9) |
| 45–64 | 1440 | (36.8) | 1404 | (38.5) | 225 | (16.0) | 178 | (12.7) | 72 | (5.2) | 27 | (1.9) | 347 | (24.7) |
| 65+ | 1454 | (37.2) | 766 | (21.0) | 66 | (8.6) | 56 | (7.3) | 19 | (2.5) | 12 | (1.6) | 112 | (14.6) |
| **Race/ethnicity§** |  |  |  |  |  | ‡ |  |  |  |  |  |  |  | ‡ |
| White, non-Hispanic | 2813 | (71.9) | 2584 | (70.8) | 479 | (18.5) | 369 | (14.3) | 131 | (5.1) | 63 | (2.4) | 684 | (26.5) |
| Black, non-Hispanic | 311 | (8.0) | 355 | (9.7) | 98 | (27.6) | 100 | (28.1) | 38 | (10.7) | 24 | (6.7) | 151 | (42.4) |
| Asian, non-Hispanic | 420 | (10.7) | 184 | (5.1) | 30 | (16.5) | 31 | (16.8) | 10 | (5.5) | 7 | (3.8) | 52 | (28.2) |
| Other race or multiple races, non-Hispanic | 116 | (3.0) | 101 | (2.8) | 31 | (31.1) | 27 | (26.4) | 4 | (3.9) | 5 | (4.9) | 44 | (43.6) |
| Hispanic, any race(s) | 251 | (6.4) | 424 | (11.6) | 146 | (34.4) | 127 | (29.9) | 48 | (11.2) | 36 | (8.6) | 200 | (47.1) |
| **Disability status\*\*** |  |  |  |  |  | ‡ |  |  |  |  |  |  |  | ‡ |
| Yes | 687 | (17.6) | 619 | (17.0) | 241 | (38.9) | 173 | (27.9) | 62 | (10.1) | 49 | (8.0) | 293 | (47.3) |
| No | 3224 | (82.4) | 3030 | (83.0) | 544 | (18.0) | 481 | (15.9) | 169 | (5.6) | 86 | (2.8) | 837 | (27.6) |
| **Marital status** |  |  |  |  |  | ‡ |  |  |  |  |  |  |  | ‡ |
| Married or living with partner | 2404 | (61.5) | 2162 | (59.2) | 366 | (16.9) | 330 | (15.3) | 106 | (4.9) | 52 | (2.4) | 563 | (26.0) |
| Divorced or separated | 462 | (11.8) | 369 | (10.1) | 73 | (19.8) | 51 | (13.9) | 25 | (6.7) | 15 | (4.2) | 105 | (28.5) |
| Never married | 825 | (21.1) | 971 | (26.6) | 314 | (32.4) | 248 | (25.5) | 90 | (9.3) | 61 | (6.3) | 422 | (43.4) |
| Widowed/widower | 220 | (5.6) | 148 | (4.1) | 31 | (20.9) | 25 | (16.6) | 10 | (6.8) | 7 | (4.7) | 41 | (27.6) |
| **Household occupancy** |  |  |  |  |  | ‡ |  |  |  |  |  |  |  | ‡ |
| Spouse or partner only | 1497 | (38.3) | 1184 | (32.4) | 173 | (14.7) | 157 | (13.3) | 56 | (4.8) | 29 | (2.4) | 277 | (23.4) |
| Child or children only | 134 | (3.4) | 130 | (3.6) | 37 | (28.5) | 25 | (19.5) | 12 | (9.5) | 5 | (4.0) | 47 | (35.9) |
| Spouse or partner and child or children | 723 | (18.5) | 819 | (22.4) | 167 | (20.4) | 138 | (16.9) | 47 | (5.7) | 20 | (2.5) | 234 | (28.5) |
| Other family member(s) only | 181 | (4.6) | 223 | (6.1) | 89 | (40.0) | 53 | (23.9) | 25 | (11.2) | 22 | (9.8) | 115 | (51.5) |
| Unrelated roommate(s) only | 44 | (1.1) | 49 | (1.3) | 17 | (35.2) | 15 | (30.8) | 6 | (11.3) | 1 | (2.8) | 25 | (51.4) |
| Pet(s) only | 301 | (7.7) | 280 | (7.7) | 71 | (25.4) | 57 | (20.5) | 10 | (3.6) | 8 | (2.9) | 94 | (33.6) |
| Other combination | 349 | (8.9) | 361 | (9.9) | 108 | (30.0) | 92 | (25.5) | 36 | (10.1) | 29 | (8.0) | 155 | (42.9) |
| No person or animal | 682 | (17.4) | 605 | (16.6) | 121 | (20.1) | 115 | (18.9) | 39 | (6.5) | 21 | (3.5) | 184 | (30.4) |
| **2019 household income (USD)** |  |  |  |  |  | ‡ |  |  |  |  |  |  |  | ‡ |
| <25,000 | 500 | (12.8) | 514 | (14.1) | 176 | (34.1) | 136 | (26.5) | 47 | (9.1) | 30 | (5.9) | 225 | (43.8) |
| 25,000–49,999 | 776 | (19.8) | 733 | (20.1) | 188 | (25.7) | 146 | (19.9) | 44 | (6.0) | 40 | (5.4) | 262 | (35.8) |
| 50,000–99,999 | 1346 | (34.4) | 1235 | (33.9) | 250 | (20.2) | 208 | (16.8) | 72 | (5.8) | 44 | (3.6) | 352 | (28.5) |
| ≥100,000 | 1289 | (33.0) | 1167 | (32.0) | 171 | (14.7) | 163 | (14.0) | 69 | (5.9) | 21 | (1.8) | 291 | (25.0) |
| **US Census region††** |  |  |  |  |  |  |  |  |  |  |  |  |  |  |
| Northeast | 897 | (22.9) | 847 | (23.2) | 175 | (20.6) | 134 | (15.9) | 50 | (5.9) | 31 | (3.6) | 245 | (28.9) |
| Midwest | 782 | (20.0) | 735 | (20.1) | 163 | (22.1) | 133 | (18.2) | 42 | (5.7) | 26 | (3.5) | 221 | (30.1) |
| South | 1228 | (31.4) | 1188 | (32.5) | 261 | (22.0) | 229 | (19.3) | 81 | (6.8) | 44 | (3.7) | 377 | (31.7) |
| West | 1004 | (25.7) | 879 | (24.1) | 186 | (21.2) | 157 | (17.8) | 59 | (6.7) | 35 | (4.0) | 287 | (32.7) |
| **Urban/rural classification‡‡** |  |  |  |  |  |  |  |  |  |  |  |  |  |  |
| Urban | 3448 | (88.2) | 3208 | (87.9) | 685 | (21.4) | 567 | (17.7) | 211 | (6.6) | 112 | (3.5) | 990 | (30.9) |
| Rural | 463 | (11.8) | 441 | (12.1) | 99 | (22.5) | 86 | (19.6) | 20 | (4.6) | 23 | (5.3) | 140 | (31.8) |
| **Employment status** |  |  |  |  |  | ‡ |  |  |  |  |  |  |  | ‡ |
| Employed | 1851 | (47.3) | 2051 | (56.2) | 455 | (22.2) | 436 | (21.2) | 154 | (7.5) | 86 | (4.2) | 686 | (33.5) |
| Retired | 1533 | (39.2) | 991 | (27.1) | 109 | (11.0) | 91 | (9.2) | 33 | (3.4) | 20 | (2.0) | 175 | (17.7) |
| Unemployed | 449 | (11.5) | 503 | (13.8) | 177 | (35.1) | 96 | (19.1) | 30 | (6.0) | 20 | (3.9) | 214 | (42.6) |
| Student | 78 | (2.0) | 104 | (2.9) | 44 | (42.0) | 30 | (29.2) | 14 | (13.4) | 10 | (9.7) | 54 | (51.6) |
| **Essential worker** |  |  |  |  |  |  |  |  |  |  |  |  |  |  |
| Yes | 842 | (45.5) | 981 | (47.8) | 225 | (22.9) | 223 | (22.8) | 86 | (8.7) | 50 | (5.1) | 345 | (35.2) |
| No | 1009 | (54.5) | 1070 | (52.2) | 230 | (21.5) | 212 | (19.9) | 68 | (6.3) | 35 | (3.3) | 341 | (31.9) |
| **Hours of paid work in previous week** |  |  |  |  |  |  |  |  |  |  |  |  |  |  |
| ≤20 | 350 | (18.9) | 344 | (16.8) | 69 | (20.1) | 63 | (18.4) | 22 | (6.4) | 13 | (3.8) | 109 | (31.7) |
| 21–40 | 1066 | (57.6) | 1201 | (58.5) | 269 | (22.4) | 261 | (21.8) | 92 | (7.7) | 53 | (4.4) | 411 | (34.2) |
| 41–60 | 389 | (21.0) | 450 | (22.0) | 99 | (22.0) | 91 | (20.2) | 31 | (6.9) | 17 | (3.8) | 142 | (31.4) |
| >60 | 46 | (2.5) | 56 | (2.7) | 18 | (32.3) | 20 | (36.3) | 8 | (14.5) | 2 | (2.9) | 25 | (44.6) |

\* See Table 2 for the adverse mental and behavioural health symptoms among those who are unpaid caregivers for adults, by select respondent demographics.
† p<0.05 for Bonferroni-corrected Rao-Scott adjusted Pearson chi-squared test between caregivers and non-caregivers.
‡ p<0.05 for Bonferroni-corrected Rao-Scott adjusted Pearson chi-squared test between demographics among caregivers.
§ “Other” race includes American Indian or Alaska Native, Native Hawaiian or Pacific Islander, or Other.
\*\* Persons who had a disability were defined as such based on a qualifying response to either one of two questions:
“Are you limited in any way in any activities because of physical, mental, or emotional condition?” and “Do you have any health conditions that require you to use special equipment, such as a cane, wheelchair, special bed, or special telephone?” https://www.cdc.gov/brfss/questionnaires/pdf-ques/2015-brfss-questionnaire-12-29-14.pdf.
†† Region classification was determined by using the US Census Bureau’s Census Regions and Divisions.
https://www2.census.gov/geo/pdfs/maps-data/maps/reference/us_regdiv.pdf.
‡‡ Rural-urban classification was determined by using self-reported ZIP codes according to the Federal Office of Rural Health Policy definition of rurality. https://www.hrsa.gov/rural-health/about-us/definition/datafiles.html.

Among caregivers, adverse mental and behavioural health symptoms were most prevalent among those aged 18–24 years (e.g., at least one symptom, *vs* those aged ≥65 years; 88.5% *vs* 18.8%, group p<0.0001), and were more prevalent among Black and Hispanic caregivers than White caregivers (80.2% and 89.4%, respectively, *vs* 53.4%, group p<0.0001) and among those with than those without disabilities (85.8% *vs* 59.8%, p<0.0001) (Table 2). There were also differences by employment status, as caregivers who were employed (76.1%) or students (79.0%) had higher prevalences of adverse mental and behavioural health symptoms than those who were retired (29.9%) or unemployed (59.3%) (group p<0.0001). Among employed caregivers, adverse mental and behavioural health symptoms were more common among essential than nonessential workers (81.6% *vs* 60.6%, p<0.0001), and were most prevalent among those who worked >60 hours in the previous week and decreased with weekly work hours (e.g., *vs* those who worked ≤20 hours; 96.9% *vs* 59.3%, group p<0.0001). There were also differences by caregiving characteristics; 93.0% of 126 caregivers providing care to multiple types of relationships reported adverse mental or behavioural health symptoms, compared with 55.6% of 261 caregivers providing care for a parent or parent-in-law (group p<0.0001). Similarly, 89.0% of 370 who had been providing care for 4–6 months, compared with 44.7% of 199 caregivers who had been providing care for more than 12 months (group p<0.0001) (Table 4). There were also difference by CII score; 91.1% of 335 caregivers with high CII scores reported at least one adverse mental or behavioural health symptom, compared with 20.7% of 31 caregivers with low CII scores (group p<0.0001).

**Table 4.**
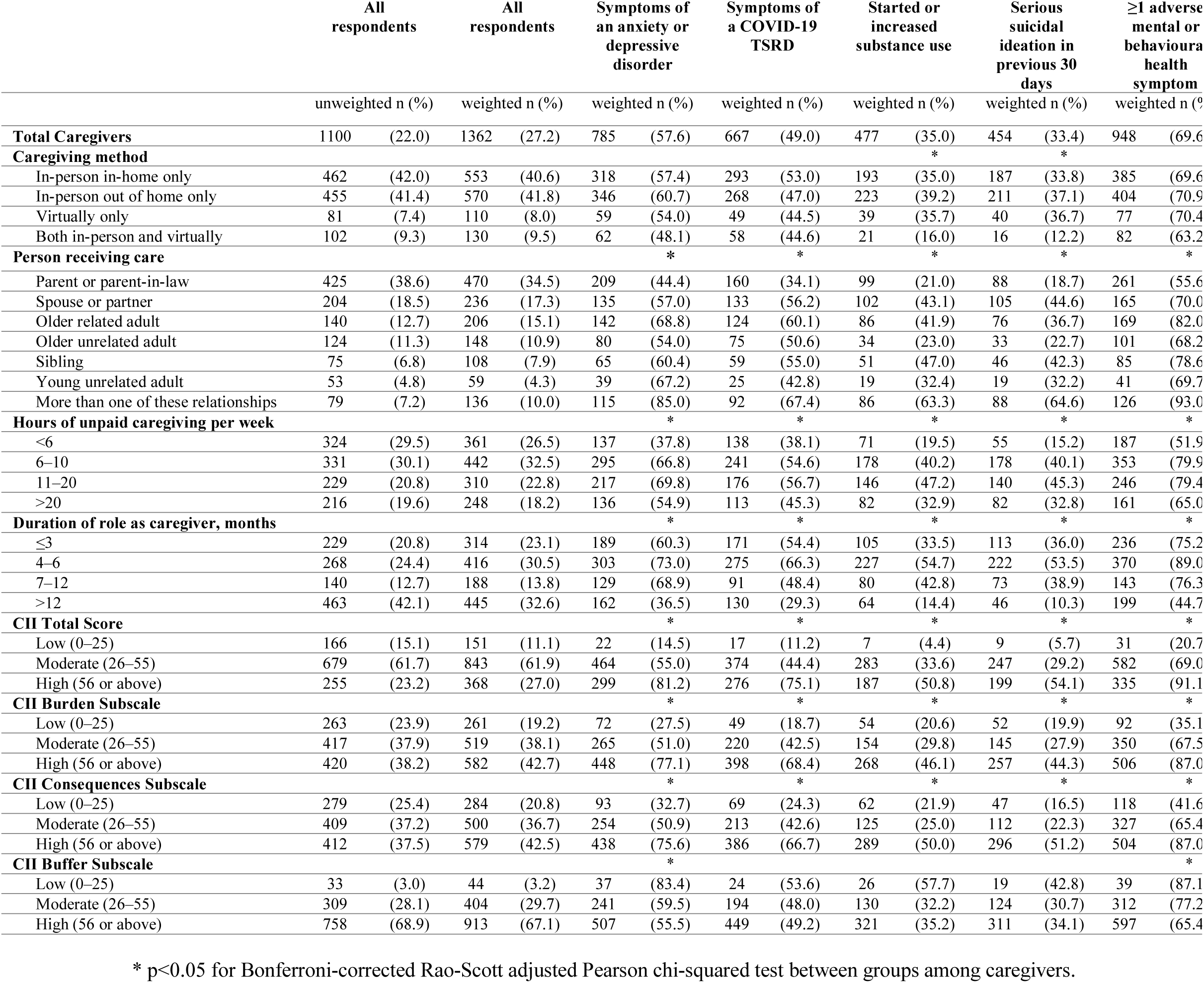
Adverse Mental and Behavioural Health Symptoms Among Unpaid Caregivers for Adults During June 24–30, 2020, by Caregiving Responsibilities and Intensity.

Adjusted prevalence ratios for select demographic and caregiving variables associated with significantly different prevalences of symptoms of anxiety or depressive disorder, suicidal ideation, and at least one adverse mental or behavioural health symptom, are shown in Figure 2. Among demographic variables, adjusted prevalence of adverse mental health symptoms was higher among young caregivers aged 18–24 years *vs* caregivers aged 45–64 years (e.g., anxiety or depressive disorder symptoms, aPR 1.47, 95% CI 1.21–1.79, p=0.0001; suicidal ideation, 1.88, 1.26–2.82, p=0.0023; at least one of these symptoms, 1.48, 1.28–1.71, p<0.0001) and those with *vs* without disabilities (1.22, 1.10–1.35, p=0.0002; 2.01, 1.65–2.46, p<0.0001; 1.18, 1.10– 1.26, p<0.0001, respectively). Suicidal ideation was more prevalent among Black *vs* White caregivers (1.48, 1.15–1.90, p=0.0022), as was at least one of these symptoms among Hispanic *vs* White caregivers (1.14, 1.04–1.25, p=0.0044). Conversely, adjusted prevalence of adverse mental health symptoms was significantly lower among older adults aged ≥65 years *vs* caregivers aged 45–64 years (e.g., at least one adverse mental health symptom, 0.54, 0.39–0.74, p=0.0002).

**Figure 2.**
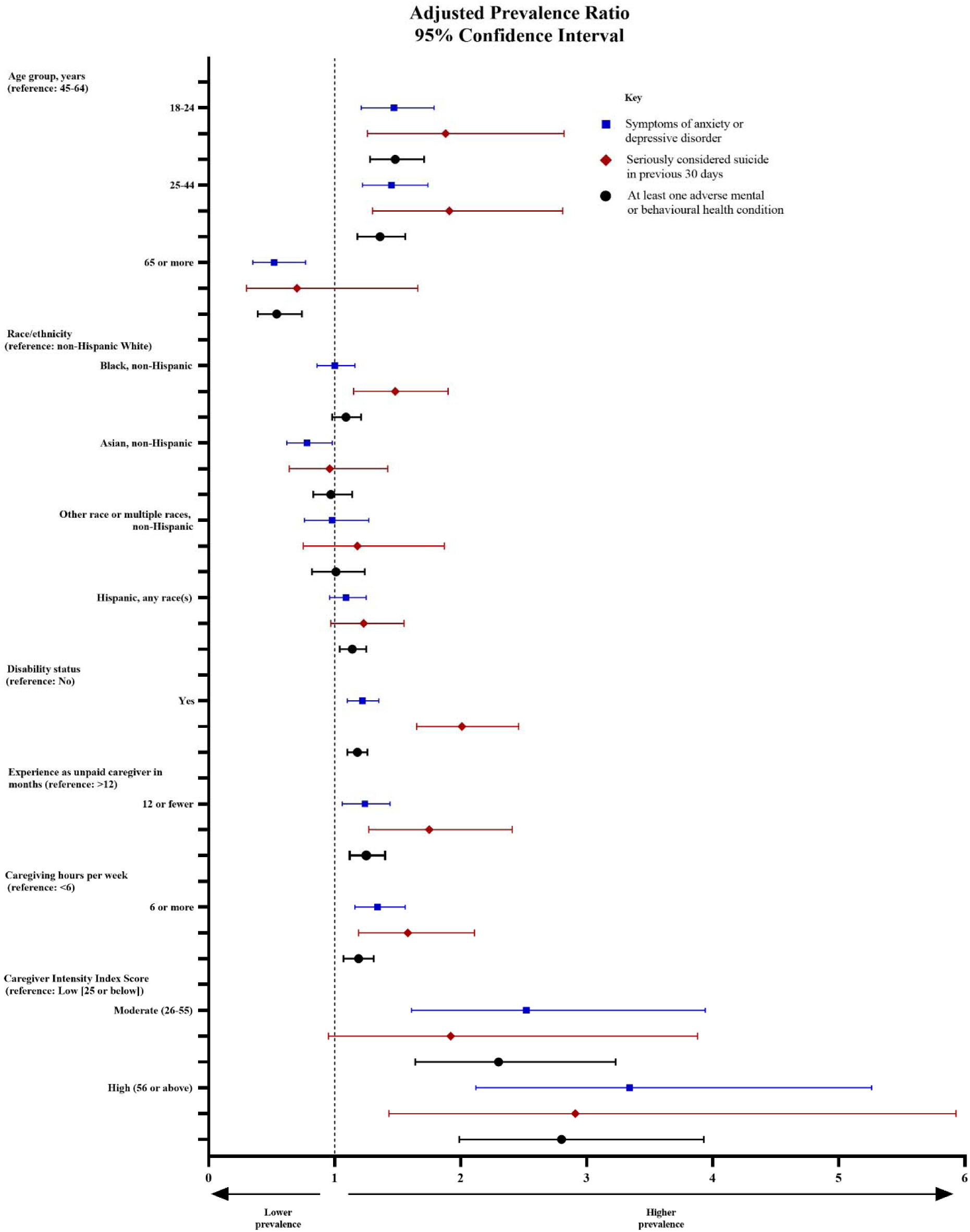
Adjusted Prevalence Ratios for Having Experienced Adverse Mental or Behavioural Health Symptoms Among Unpaid Caregivers for Adults, by Select Respondent Characteristics.

Among caregiving variables, adjusted prevalence of adverse mental health symptoms was higher among caregivers with ≤12 *vs* those with >12 months of experience (anxiety or depressive disorder symptoms, 1.24, 1.06–1.44, p=0.0059; suicidal ideation, 1.75, 1.27–2.41, p=0.0006; at least one of these symptoms, 1.25, 1.12–1.40, p=0.0001), those with >6-*vs* ≤6-hour weekly caregiving commitment (1.34, 1.16–1.56, p=0.0001; 1.58, 1.19–2.11, p=0.0018; 1.19, 1.07–1.31, p=0.0009, respectively), and, compared with those in the low-intensity CII group, those in the moderate-intensity (2.52, 1.61–3.94, p<0.0001; 1.92, 0.95–3.88, p=0.070; 2.30, 1.64–3.23, p<0.0001, respectively) and high-intensity (3.34, 2.12–5.26, p<0.0001; 2.91, 1.43–5.93, p=0.0034; 2.80, 1.99–3.93, p<0.0001, respectively) groups.

In the exploratory analysis of the correlation of individual CII items with adverse mental and behavioural health symptoms, the strongest average positive correlations among all adverse symptoms were observed for employment absenteeism (ρs between 0.36 and 0.46, all p<0.0001), preparedness (ρs between 0.25 and 0.45, all p <0.0001), resentment (ρs between 0.30 and 0.40, all p<0.0001), impact on expenses (ρs between 0.26 and 0.45, all p <0.0001), and family strife (ρs between 0.24 and 0.42, all p<0.0001) (Table 5). The strongest average negative correlation was observed for sense of purpose (ρs between -0.11 and -0.22, all p ≤0.0002). All correlations were in the expected direction based on their subscale categorization, except for employer support, which had a positive correlation with all adverse mental or behavioural health symptoms (ρs between 0.16 and 0.26, all p <0.0001) despite being in the Buffer subscale.

**Table 5.**
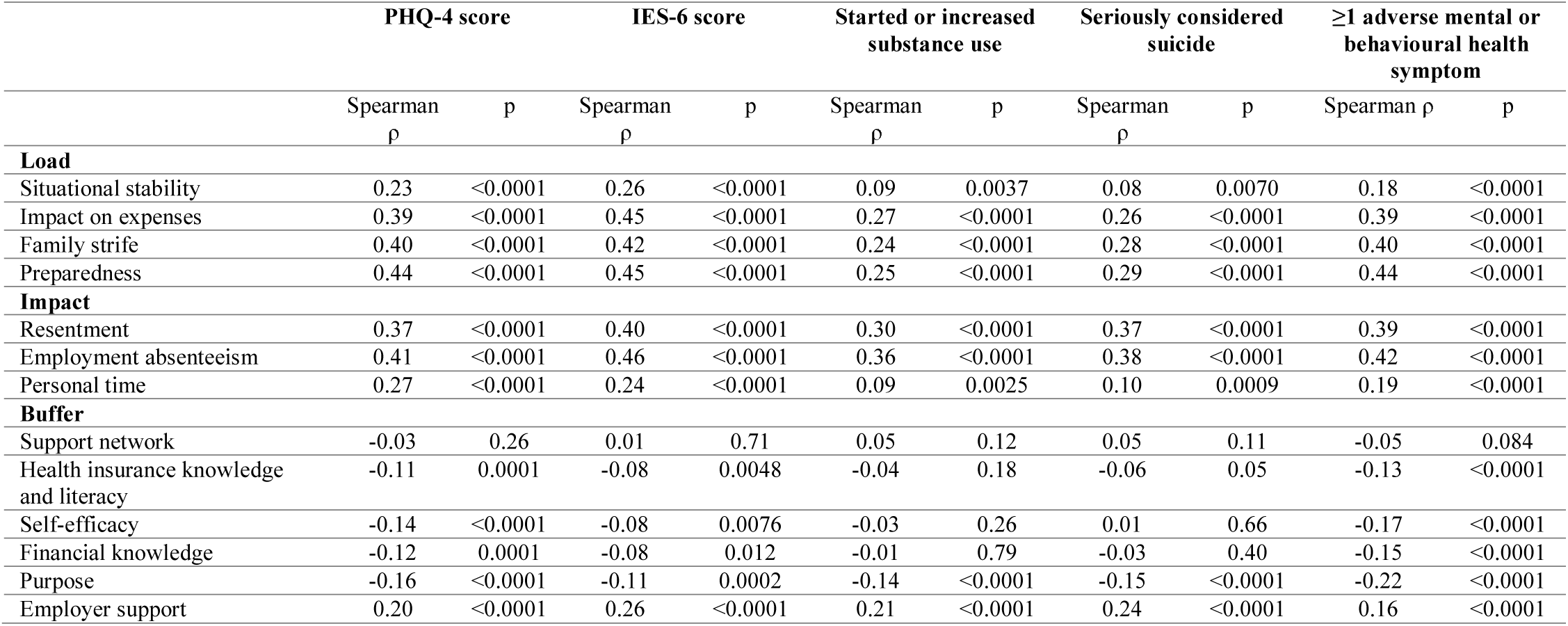
Correlation Between Individual Caregiver Intensity Index Items and Adverse Mental or Behavioural Health Symptoms.

|  | PHQ-4 score |  | IES-6 score |  | Started or increased substance use |  | Seriously considered suicide |  | ≥1 adverse mental or behavioural health symptom |  |
| --- | --- | --- | --- | --- | --- | --- | --- | --- | --- | --- |
| | Spearman $\rho$ | p | Spearman $\rho$ | p | Spearman $\rho$ | p | Spearman $\rho$ | p | Spearman $\rho$ | p |
| **Load** |  |  |  |  |  |  |  |  |  |  |
| Situational stability | 0.23 | <0.0001 | 0.26 | <0.0001 | 0.09 | 0.0037 | 0.08 | 0.0070 | 0.18 | <0.0001 |
| Impact on expenses | 0.39 | <0.0001 | 0.45 | <0.0001 | 0.27 | <0.0001 | 0.26 | <0.0001 | 0.39 | <0.0001 |
| Family strife | 0.40 | <0.0001 | 0.42 | <0.0001 | 0.24 | <0.0001 | 0.28 | <0.0001 | 0.40 | <0.0001 |
| Preparedness | 0.44 | <0.0001 | 0.45 | <0.0001 | 0.25 | <0.0001 | 0.29 | <0.0001 | 0.44 | <0.0001 |
| **Impact** |  |  |  |  |  |  |  |  |  |  |
| Resentment | 0.37 | <0.0001 | 0.40 | <0.0001 | 0.30 | <0.0001 | 0.37 | <0.0001 | 0.39 | <0.0001 |
| Employment absenteeism | 0.41 | <0.0001 | 0.46 | <0.0001 | 0.36 | <0.0001 | 0.38 | <0.0001 | 0.42 | <0.0001 |
| Personal time | 0.27 | <0.0001 | 0.24 | <0.0001 | 0.09 | 0.0025 | 0.10 | 0.0009 | 0.19 | <0.0001 |
| **Buffer** |  |  |  |  |  |  |  |  |  |  |
| Support network | -0.03 | 0.26 | 0.01 | 0.71 | 0.05 | 0.12 | 0.05 | 0.11 | -0.05 | 0.084 |
| Health insurance knowledge and literacy | -0.11 | 0.0001 | -0.08 | 0.0048 | -0.04 | 0.18 | -0.06 | 0.05 | -0.13 | <0.0001 |
| Self-efficacy | -0.14 | <0.0001 | -0.08 | 0.0076 | -0.03 | 0.26 | 0.01 | 0.66 | -0.17 | <0.0001 |
| Financial knowledge | -0.12 | 0.0001 | -0.08 | 0.012 | -0.01 | 0.79 | -0.03 | 0.40 | -0.15 | <0.0001 |
| Purpose | -0.16 | <0.0001 | -0.11 | 0.0002 | -0.14 | <0.0001 | -0.15 | <0.0001 | -0.22 | <0.0001 |
| Employer support | 0.20 | <0.0001 | 0.26 | <0.0001 | 0.21 | <0.0001 | 0.24 | <0.0001 | 0.16 | <0.0001 |

## Discussion

More than one quarter (1,362 [27.2%]) of 5,011 US adult respondents identified as unpaid caregivers for adults in the three months preceding the survey in June 2020. This estimated prevalence of caregivers in the United States during the COVID-19 pandemic represents an increase over the 19.2% estimate based on data collected in 2019,^9^ which may reflect an impact of the pandemic on caregivers and recipients of care. Overall, 7 in 10 (948 of 1,362 [69.6%]) caregivers reported having experienced at least one adverse mental or behavioural health symptom. More than half of caregivers screened positive for symptoms of an anxiety or depressive disorder (785 [57.2%]), and more than one-third reported having started or increased substance use to cope with the stress or emotions related to COVID-19 (477 [35.0%]) and seriously considered suicide in the prior month (454 [33.4%]). Caregivers reported having experienced elevated levels of adverse mental and behavioural health symptoms compared to non-caregivers in this study, including three-fold increased prevalences of symptoms of anxiety or depressive disorder or a COVID-19-related TSRD, six-fold increased prevalence of having started or increased substance use to cope with the pandemic, and nine-fold increased prevalence of caregivers having seriously considered suicide compared to non-caregivers.

Both caregivers and non-caregivers who were young, Black, Hispanic, living with disabilities, essential workers, and working long hours had disproportionately high levels of adverse mental health, consistent with findings during the pandemic.^13-16^ However, caregivers more commonly identified as members of these at-risk populations than non-caregivers, with more than two-thirds (928 [68.1%]) of caregivers aged below 45 years, more than half (756 [55.5%]) non-White, more than one-third living with disabilities (516 [37.9%]), and nearly three-quarters of caregivers employed as essential workers (751 of 1,018 [73.7%]), adding additional potential stressors to their caregiving responsibilities. Long work hours, which were also common among employed caregivers, are associated with increased odds of adverse health outcomes, including depression, anxiety, and impaired sleep,^17^ an effect that may be exacerbated by caregiving responsibilities outside of work. Committing long hours to paid work and unpaid care limits opportunities for core elements of health, including sleeping, exercising, eating, socializing, and medical care. Among caregivers, those who provide care for more hours and those who had been caregiving for fewer than 12 months had higher prevalences of adverse mental health symptoms, which may reflect stressors from being forced into a caregiving role, starting as a caregiver during the pandemic, or survival bias, whereby those who are still providing care after 12 months are more resilient to stressors associated with the role.

The findings in this report reveal that unpaid caregiving for adults is common, has likely increased during the COVID-19 pandemic, and is represented broadly across demographics. Further, the report underscores the significant impact associated with caregiving on mental and behavioural health and highlights the compounding impact of intersectionality with those who identify in multiple groups having elevated experiences of adverse mental and behavioural health. Addressing mental health among caregivers represents an urgent unmet medical need, and targeted interventions and communication strategies are needed to increase awareness of, comfort with, and access to resources for diagnosis and treatment of mental and behavioural health symptoms, especially given the time constraints faced by caregivers, many of whom are also employed. Effective communication strategies may include promoting recognition of caregivers so that they feel seen,^18^ addressing stigma associated with mental healthcare,^19-21^ along with continuing to expand telehealth,^22^ which has delivered promising results in treatment for mental health conditions, including depression, substance use disorder, and suicidal ideation.^23^ However, telehealth may not address all needs, with barriers to access (i.e., lack of Internet access) and limitations to provision of some care. Campaigns to increase help-seeking behaviour may also be beneficial, as caregivers more commonly avoided medical care due to concerns about COVID-19,^24^ which may be related to their own perceived risk of SARS-CoV-2 infection, to their perceived risk and grief about potentially infecting the person for whom they are caring, or both.

The COVID-19 pandemic both introduced new challenges (e.g., barriers to in-person care provision, COVID-19 concerns) and exacerbated the challenges (e.g., financial strains) associated with caregiving that existed before the pandemic. Therefore, prevention efforts and cultural changes may be required both during and beyond the pandemic to properly address the factors associated with caregiving that contribute to elevated experiences of adverse mental health. This is of increasing importance to the economy, as even before the expected impact of COVID-19, a 2015 study estimated the value of unpaid caregiver labour to be USD$470 billion.^25^ It is also important to note that the impact of caregiving does not end with the death of the loved ones. Pre-pandemic research highlighted that approximately 20% of bereaved caregivers experience psychiatric symptoms, including depression and/or complicated grief following the passing of their loved ones.^26^ Given the high prevalence of employed caregivers and its compounding mental health impact, reducing the stigma that can be associated with caregiver status and establishing visible and easily accessible workplace programs should be prioritized. Employee Assistance Programs, Workplace Health Promotion Programs, personalized flexible work arrangements, and expanded options for leave that may reduce caregiving intensity if effectively utilized.^27^ Assistive technologies may also decrease workloads required from caregivers, though may inadvertently increase the load if mismanaged or improperly designed.^28^ Beyond these institutional changes, given the protective benefit of a caregiver’s sense of purpose and evidence that self-esteem and positive aspects of caregiving are associated with improved mental health,^29^ creating a culture that more openly celebrates caregivers and their efforts may lead to communities of caregivers that reduce the mental health risks associated with social disconnectedness and isolation.^30-32^

Strengths of this study include recruitment of a large sample of unpaid caregivers from a nationally representative sample of U.S. adults and utilization of validated screening instruments for mental health. This paper is subject limitations, which may include the following. First, unpaid caregivers for adults were self-identified, and caregiver status for children or adolescents were not assessed; future research should assess mental health among multigenerational caregivers. Second, a diagnostic evaluation for anxiety disorder or depressive disorder was not conducted; however, clinically validated screening instruments were used to assess symptoms. Third, substance use was self-reported; therefore, responses might be subject to recall, response, and social desirability biases. Fourth, the novel nature of the ARCHANGELS Caregiver Intensity Index and the specific use within this research precludes exact comparisons with normative data on caregiving intensity before the pandemic. Finally, the Internet-based survey may not be fully representative of the 2020 U.S. population and may therefore have limited generalizability. However, standardized and supplementary quality and data inclusion screening procedures were applied, and the prevalence of symptoms of anxiety disorder and depressive disorder were largely consistent with findings from the Household Pulse Survey during June.^14^

Further characterization of caregivers and assessment of mental health, substance use, and suicidal ideation will be required to determine the extent to which increased prevalence of caregiving and elevated adverse mental and behavioural health symptoms progress over the course of the pandemic and beyond. Investment in support systems that reflect the diverse caregiving population and improves their ability to provide care will improve societal health and well-being during this critical health crisis and beyond.

## Data Availability

All relevant data are presented in the report and available from the corresponding author upon reasonable request.

## Contributors

All authors contributed to the study concept and design. MÉC, CAC, SMWR, and MEH collected the data, and MÉC conducted all analyses and wrote the first manuscript draft. All authors provided critical intellectual input and revision. SMWR and MEH provided supervision. MÉC, SMWR, and MEH had access to the underlying data.

## Acknowledgements

The authors gratefully acknowledge all survey respondents, along with Elise R Facer-Childs, PhD, Monash University, Rebecca Robbins, PhD, Brigham & Women’s Hospital, and Laura K Barger, PhD, Brigham & Women’s Hospital, for their contributions to the survey instrument and study design. The authors also thank Mallory Colys, Sneha Baste, Daniel Chong, and Rebecca Toll, Qualtrics LLC, for their support in the survey administration, and Wendy D Lynch, PhD, Lynch Consulting, Ltd, Indiana University School of Nursing, for assistance with deploying an abbreviated form of the ARCHANGELS Caregiver Intensity Index.

## References

1. Czeisler MÉ, Lane RI, Petrosky E, et al. Mental Health, Substance Use, and Suicidal Ideation During the COVID-19 Pandemic - United States, June 24-30, 2020. MMWR Morb Mortal Wkly Rep 2020; 69(32): 1049–57.

2. Ettman CK, Abdalla SM, Cohen GH, Sampson L, Vivier PM, Galea S. Prevalence of Depression Symptoms in US Adults Before and During the COVID-19 Pandemic. JAMA Netw Open 2020; 3(9): e2019686.

3. Pierce M, Hope H, Ford T, et al. Mental health before and during the COVID-19 pandemic: a longitudinal probability sample survey of the UK population. Lancet Psychiatry 2020; 7(10): 883–92.

4. Shi L, Lu ZA, Que JY, et al. Prevalence of and Risk Factors Associated With Mental Health Symptoms Among the General Population in China During the Coronavirus Disease 2019 Pandemic. JAMA Netw Open 2020; 3(7): e2014053.

5. Pinquart M, Sorensen S. Differences between caregivers and noncaregivers in psychological health and physical health: a meta-analysis. Psychol Aging 2003; 18(2): 250–67.

6. Del-Pino-Casado R, Rodriguez Cardosa M, Lopez-Martinez C, Orgeta V. The association between subjective caregiver burden and depressive symptoms in carers of older relatives: A systematic review and meta-analysis. PLoS One 2019; 14(5): e0217648.

7. Fekete C, Tough H, Siegrist J, Brinkhof MW. Health impact of objective burden, subjective burden and positive aspects of caregiving: an observational study among caregivers in Switzerland. BMJ Open 2017; 7(12): e017369.

8. Voisin MR, Oliver K, Farrimond S, et al. Brain tumors and COVID-19: the patient and caregiver experience. Neurooncol Adv 2020; 2(1): vdaa104.

9. Caregiving in the US: 2020 report. Washington, D.C.: National Alliance for Caregiving; AARP Public Policy Institute; 2020.

10. Caregiving in the US: 2015 report. National Alliance for Caregiving; AARP Public Policy Institute; 2015.

11. Lowe B, Wahl I, Rose M, et al. A 4-item measure of depression and anxiety: validation and standardization of the Patient Health Questionnaire-4 (PHQ-4) in the general population. J Affect Disord 2010; 122(1-2): 86–95.

12. Hosey MM, Leoutsakos JS, Li X, et al. Screening for posttraumatic stress disorder in ARDS survivors: validation of the Impact of Event Scale-6 (IES-6). Crit Care 2019; 23(1): 276.

13. Gold JA. Covid-19: adverse mental health outcomes for healthcare workers. BMJ 2020; 369: m1815.

14. CDC NCfHS. Indicators of anxiety or depression based on reported frequency of symptoms during the last 7 days. Household Pulse Survey. In: Services UDoHaH, editor. Atlanta, GA: CDC, National Center for Health Statistics; 2020.

15. Wang X, Hegde S, Son C, Keller B, Smith A, Sasangohar F. Investigating Mental Health of US College Students During the COVID-19 Pandemic: Cross-Sectional Survey Study. J Med Internet Res 2020; 22(9): e22817.

16. Son C, Hegde S, Smith A, Wang X, Sasangohar F. Effects of COVID-19 on College Students’ Mental Health in the United States: Interview Survey Study. J Med Internet Res 2020; 22(9): e21279.

17. Wong K, Chan AHS, Ngan SC. The Effect of Long Working Hours and Overtime on Occupational Health: A Meta-Analysis of Evidence from 1998 to 2018. Int J Environ Res Public Health 2019; 16(12).

18. O’connor DL. Self-identifying as a caregiver: Exploring the positioning process. Journal of Aging Studies 2007; 21(2): 165–74.

19. Picco L, Abdin E, Pang S, et al. Association between recognition and help-seeking preferences and stigma towards people with mental illness. Epidemiol Psychiatr Sci 2018; 27(1): 84–93.

20. Schomerus G, Stolzenburg S, Freitag S, et al. Stigma as a barrier to recognizing personal mental illness and seeking help: a prospective study among untreated persons with mental illness. Eur Arch Psychiatry Clin Neurosci 2019; 269(4): 469–79.

21. Horsfield P, Stolzenburg S, Hahm S, et al. Self-labeling as having a mental or physical illness: the effects of stigma and implications for help-seeking. Soc Psychiatry Psychiatr Epidemiol 2020; 55(7): 907–16.

22. Koonin LM, Hoots B, Tsang CA, et al. Trends in the Use of Telehealth During the Emergence of the COVID-19 Pandemic - United States, January-March 2020. MMWR Morb Mortal Wkly Rep 2020; 69(43): 1595–9.

23. Hailey D, Roine R, Ohinmaa A. The effectiveness of telemental health applications: a review. Can J Psychiatry 2008; 53(11): 769–78.

24. Czeisler MÉ, Marynak K, Clarke KEN, et al. Delay or Avoidance of Medical Care Because of COVID-19-Related Concerns - United States, June 2020. MMWR Morb Mortal Wkly Rep 2020; 69(36): 1250–7.

25. Beltran-Sanchez H, Soneji S, Crimmins EM. Past, Present, and Future of Healthy Life Expectancy. Cold Spring Harb Perspect Med 2015; 5(11).

26. Schulz R, Hebert R, Boerner K. Bereavement after caregiving. Geriatrics 2008; 63(1): 20–2.

27. Lilly MB. The hard work of balancing employment and caregiving: what can canadian employers do to help? Healthc Policy 2011; 7(2): 23–31.

28. Marasinghe KM, Lapitan JM, Ross A. Assistive technologies for ageing populations in six low-income and middle-income countries: a systematic review. BMJ Innov 2015; 1(4): 182–95.

29. Fauziana R, Sambasivam R, Vaingankar JA, et al. Positive Caregiving Characteristics as a Mediator of Caregiving Burden and Satisfaction With Life in Caregivers of Older Adults. J Geriatr Psychiatry Neurol 2018; 31(6): 329–35.

30. Newman MG, Zainal NH. The value of maintaining social connections for mental health in older people. Lancet Public Health 2020; 5(1): e12–e3.

31. Hammig O. Health risks associated with social isolation in general and in young, middle and old age. PLoS One 2019; 14(7): e0219663.

32. Bhatti AB, Haq AU. The Pathophysiology of Perceived Social Isolation: Effects on Health and Mortality. Cureus 2017; 9(1): e994.

