## Supplement for "Mental Health, Substance Use, and Suicidal Ideation Among Unpaid Caregivers in the United States During the COVID-19 Pandemic: Relationships to Age, Race/Ethnicity, Employment, and Caregiver Intensity"

In this section, we provide more details about the Qualtrics, LLC, (henceforth, Qualtrics) the data source, and about its advantages and limitations.

### Recruitment

**Qualtrics LLC methodology**
Qualtrics partners with sample partners that employ various recruitment methodologies, with most samples come from traditional, actively managed, double-opt-in market research panels, through which sample partners randomly select respondents for surveys where respondents are likely to qualify. Most survey invitations are sent via email, in-mobile-application notifications, and Short Message Service (SMS) notifications, and do not include information about the contents of the survey to reduce self-selection bias. Respondents receive incentives, which vary and may include cash, airline miles, gift cards, redeemable points, charitable donations, sweepstakes entrance, and vouchers, with the level of incentive based on a number of factors, including the length of the survey.

For additional information, ESOMAR (www.esomar.org) provides a Guideline for Online Research that provides further detail for how Qualtrics aims for best practices in the handling of ethical, methodological, and regulatory issues, as well as the legalities regarding technology in research.

For this study, Qualtrics was selected based on its demonstrated ability to recruit more demographically representative samples compared with alternative online survey companies (eg, Facebook, Amazon Mechanical Turk).[1]

### Measures

**Caregiving intensity – ARCHANGELS Caregiving Intensity Index**
The ARCHANGELS Caregiver Intensity Index (CII) is a proprietary tool developed by ARCHANGELS (Boston, Massachusetts, USA; https://www.archangels.me/) to assess global caregiving intensity (CII Total Score), comprised of the following subscales: caregiving load (CII Load), caregiving impact (CII Impact), and caregiving buffers (CII Buffer). For this study, a reduced 14-item (employed) or 12-item (unemployed) CII was completed by respondents who self-identified as unpaid caregivers for adults. For each item, respondents indicated the extent to which they agree with statements regarding each of the topics below as they pertain to caregiving.

| |
|---|
| **Load Items:** Situational stability, Impact on expenses, Family strife, Preparedness |
| **Impact Items:** Resentment, Employment absenteeism,* Personal time |
| **Buffer Items:** Support network, Health insurance knowledge and literacy, Self-efficacy, Financial knowledge, Purpose, Employer support* |

* employed caregivers only

The investigators received permission to use the ARCHANGELS CII for research purposes at no cost.

**Mental and behavioural health screening tools**
For anxiety and depressive disorders symptoms assessed via the 4-item Patient Health Questionnaire (PHQ-4), respondents who scored ≥3 out of 6 on the 2-item Generalized Anxiety Disorder (GAD-2) or 2-item Patient Health Questionnaire (PHQ-2) subscale were considered symptomatic for an anxiety disorder or depressive disorder, respectively. For COVID-19 TSRD symptoms assessed via the 6-item Impact of Event Scale (IES-6), in this survey,

1

the COVID-19 pandemic was specified as the traumatic exposure to record peri- and posttraumatic symptoms associated with the range of stressors introduced by the COVID-19 pandemic, and respondents who scored ≥1.75 out of 4 were considered symptomatic. Substance use was defined as use of "alcohol, legal or illegal drugs, or prescriptions drugs that are taken in a way not recommended by your doctor."

For the questions about substance use and serious suicidal ideation, all respondents were provided with mental and behavioural health resources and informed that responses were anonymised and direct support could therefore not be provided.

### Quality Control

**Qualtrics LLC response screening**
Qualtrics employs multiple algorithmic analyses of survey responses to improve the quality of responses through the identification of click thru behaviour, duplicate responses, machine responses (ie, bots), inattentiveness, and through geolocation verification via country-level Internet Protocol (IP) address mapping. For this analysis, geolocation verification was employed to confirm that respondents who self-reported residence within the US were responding to the survey from within the US.

**Secondary response screening and inclusion criteria**
In addition to the Qualtrics standard measures for quality control, given the encoding of ZIP codes as variables in this analysis, supplementary cleaning of ZIP codes was conducted to ensure all manually entered values were valid US ZIP codes. Respondents who did not provide sufficient detail to characterize responses for variables included in this analysis were excluded.

### Statistical Analysis

**Survey weighting**
Iterative proportional fitting (raking) and weight trimming (0.3≤weight≤3.0) was employed using the R survey package to improve the cross-sectional sample representativeness of the 2010 US population by age, gender, and combined race/ethnicity. Population estimates from the 2010 US Census were used. For sample gender, given that the Census did not assess gender, estimates for sex were used. For combined race/ethnicity, race and ethnicity were collected as separate questions in the survey, per the Census, and combined during analysis.

### References


1. Boas TC, Christenson DP, Glick DM. Recruiting large online samples in the United States and India: Facebook, Mechanical Turk, and Qualtrics. *Political Science Research and Methods*. 2020;**8**(2):232-50.
